## Supplementary materials for "Clinician perspectives on what constitutes good practice in community services for people with Complex Emotional Needs: A qualitative thematic meta-synthesis"

**Appendix A – Search Strategy**

The purpose of this search is to identify studies reporting provision of community-based care for people with diagnosed personality disorder. The search strategy was developed by the review team and it incorporates feedback from our PPI group (the LEWG) who worked with us on this review.

The bibliographic search strategy set out below aims to identify published studies or reports. This search narrative seeks to illustrate the conceptual purpose of the search strategy and the contextual detail of the search syntax reported below (Cooper et al. 2018). Separate search strategies – based on the syntax reported below – will be devised for web-searching.

| **#** | **Searches** | **Results** | **Narrative** |
| --- | --- | --- | --- |
| 1 | exp *Personality Disorders/ | 28263 | 1. This controlled indexing term has been exploded (indicated by exp) to capture other relevant MeSH indexing terms, such as:   - Antisocial Personality Disorder/ - Borderline Personality Disorder/ - Compulsive Personality Disorder/ - Dependent Personality Disorder/ - Histrionic Personality Disorder/ - Hysteria/ - Paranoid Personality Disorder/ - Passive‐Aggressive Personality Disorder/ - Schizoid Personality Disorder/ - Schizotypal Personality Disorder/   2. Line two picks up the controlled indexing terms as free-text terms in the title (ti), abstract (ab) and author-generated keyword (kw). A proximity marker has been included (adj3) to identify personality within two words of the other terms in the search line. These free-text terms were verified against organisations such as MIND, for completeness. These search terms have been resented at a high-level for the purposes of scoping. The aim of this searching is not comprehensive but for a realistic sample of studies and data.  4. Search terms suggested by a member of the LEWG. Terms related to CPTSD which many of those diagnosed will have, and may provide useful models of care.  5. Suggested by a member of the LEWG. Services that don’t used the personality disorder terminology often refer to themselves as “complex needs services” or “complex mental health”  6/7. Suggested by a member of the LEWG. Services for self-harm often see large numbers of people with personality disorder diagnoses.  8. Suggested by members of the LEWG.  10. combines the population-level search terms using the Boolean connect or, so that all terms represented are identified. |
| 2 | ((personality or character*) adj3 disorder$).ti,ab,kw. | 65164 |  |
| 3 | "axis II".ti,ab,kw. | 1959 |  |
| 4 | ("Complex trauma" or CPTSD or "complex post-traumatic stress disorder").ti,ab,kw. | 535 |  |
| 5 | (Complex adj (needs or mental)).ti,ab,kw. | 1929 |  |
| 6 | *Self-Injurious Behavior/ | 5465 |  |
| 7 | (Self-harm or self-injury).ti,ab,kw. | 7484 |  |
| 8 | (emotion* adj2 (regulation or dysregulation or unstable or instability)).ti,ab,kw. | 10588 |  |
| 9 | mood instability.ti,ab,kw. | 254 |  |
| 10 | 1 or 2 or 3 or 4 or 5 or 6 or 7 or 8 or 9 | 103674 |  |
| 11 | Community Health Services/ | 31050 | 11-18. These terms are for settings, including controlled indexing terms for relevant forms of health care provision and corresponding free-text terminology.  19. combines population and intervention terminology. |
| 12 | Community Mental Health Services/ | 18317 |  |
| 13 | ((commun$ adj5 (mental health or model$1 or pathway$1 or program$ or evaluat$ or intervention$ or implement$)) or camhs or cmht$1).ti,ab,kw. | 79584 |  |
| 14 | (community adj5 (agenc$ or care or center$ or centre$ or clinic$ or consultant$ or doctor$ or employee$ or expert$ or facilitator$ or healthcare or instructor$ or leader$ or manager$ or mentor$ or nurs$ or personnel$ or pharmacy or pharmacist$ or psychiatrist$ or psychologist$ or psychotherapist$ or specialist$ or skill$ or staff$ or team$ or therapist$ or tutor$ or visit$ or worker$ or group$ or independent or (peer$ adj3 support$) or survivor or outpatient$ or "out patient$")).ti,ab,kw. | 96749 |  |
| 15 | (commun$ adj5 (service or hub$ or based or deliver$ or interact$ or led or maintenance or mediat$ or operated or provides or provider$ or run or setting$ or support or rehab$ or therap$ or service$ or treatment or management or assessment or assistance or care or day or week)).ti,ab,kw. | 205151 |  |
| 16 | (Independent sector or ((non institutional$ or noninstitution$) adj2 (sector$ or setting$))).ti,ab,kw. | 367 |  |
| 17 | (network or outreach or ((specialist or day or whole) adj3 service)).ti,ab,kw. | 351098 |  |
| 18 | ((treatment* or (Dialectical behavior therapy or Dialectical behaviour therapy or DBT) or Psychotherapy* or specialist or psychiatry* or therapeutic or day or outreach or therap*) adj3 (Outpatient* or community or Service* or Center* or Centre* or Clinic*1 or Team* or program* or provider* or practice or setting* or care or community or unit* or hospital*)).ti,ab,kw. | 244595 |  |
| 19 | 11 or 12 or 13 or 14 or 15 or 16 or 17 or 18 | 851066 |  |
| 20 | Interview*.af. | 373632 | 20-24. are the Wong broad filter for Qualitative studies (Wong et al. 2004) |
| 21 | Experience*.af. | 1028333 |  |
| 22 | qualitative.tw. | 212806 |  |
| 23 | Qualitative Research/ | 50164 |  |
| 24 | 20 or 21 or 22 or 23 | 1444445 |  |
| 25 | randomized controlled trial.pt. | 495635 | 25-45. are the Cochrane HSSS (Lefebvre, Manheimer, Glanville, 2011) and the SIGN observational search filter (Scottish Intercollegiate Guidelines Network, 2018) for studies reporting controlled trials and observational studies. |
| 26 | controlled clinical trial.pt. | 93449 |  |
| 27 | (randomized or randomised).ab. | 553632 |  |
| 28 | placebo.ab. | 203251 |  |
| 29 | clinical trials as topic.sh. | 189357 |  |
| 30 | randomly.ab. | 322897 |  |
| 31 | trial.ti. | 209094 |  |
| 32 | 25 or 26 or 27 or 28 or 29 or 30 or 31 | 1289520 |  |
| 33 | Epidemiologic studies/ | 8156 |  |
| 34 | exp case control studies/ | 1037554 |  |
| 35 | Case control.tw. | 120110 |  |
| 36 | (cohort adj (study or studies)).tw. | 190004 |  |
| 37 | Cohort analy$.tw. | 7484 |  |
| 38 | (Follow up adj (study or studies)).tw. | 47969 |  |
| 39 | (observational adj (study or studies)).tw. | 98982 |  |
| 40 | Longitudinal.tw. | 232846 |  |
| 41 | Retrospective.tw. | 497909 |  |
| 42 | Cross sectional.tw. | 329612 |  |
| 43 | Cross-sectional studies/ | 311409 |  |
| 44 | 33 or 34 or 35 or 36 or 37 or 38 or 39 or 40 or 41 or 42 or 43 | 2028189 |  |
| 45 | 32 or 44 | 3197720 |  |
| 46 | "Surveys and Questionnaires"/ | 443562 | 46-47. represent a pragmatic search for studies reporting surveys. |
| 47 | survey$.tw. | 612248 |  |
| 48 | exp clinical pathway/ | 6469 | 48-69. is the CADTH search filter for guidelines (CADTH, 2018).  70 combines the search filters.  71 includes a search for systematic reviews on this topic for the supplementary searches.  72 combines the population-level terms with the community-level intervention terms and the search filters. |
| 49 | exp clinical protocol/ | 163100 |  |
| 50 | exp consensus/ | 11712 |  |
| 51 | exp consensus development conference/ | 11685 |  |
| 52 | exp consensus development conferences as topic/ | 2772 |  |
| 53 | critical pathways/ | 6469 |  |
| 54 | exp guideline/ | 33000 |  |
| 55 | guidelines as topic/ | 38818 |  |
| 56 | exp practice guideline/ | 26125 |  |
| 57 | health planning guidelines/ | 4067 |  |
| 58 | (guideline or practice guideline or consensus development conference or consensus development conference, NIH).pt. | 42151 |  |
| 59 | (position statement* or policy statement* or practice parameter* or best practice*).ti,ab,kf,kw. | 31048 |  |
| 60 | (standards or guideline or guidelines).ti,kf,kw. | 105440 |  |
| 61 | ((practice or treatment* or clinical) adj guideline*).ab. | 37832 |  |
| 62 | (CPG or CPGs).ti. | 5569 |  |
| 63 | consensus*.ti,kf,kw. | 24689 |  |
| 64 | consensus*.ab. /freq=2 | 23911 |  |
| 65 | ((critical or clinical or practice) adj2 (path or paths or pathway or pathways or protocol*)).ti,ab,kf,kw. | 19229 |  |
| 66 | recommendat*.ti,kf,kw. | 39030 |  |
| 67 | (care adj2 (standard or path or paths or pathway or pathways or map or maps or plan or plans)).ti,ab,kf,kw. | 55156 |  |
| 68 | (algorithm* adj2 (screening or examination or test or tested or testing or assessment* or diagnosis or diagnoses or diagnosed or diagnosing)).ti,ab,kf,kw. | 7192 |  |
| 69 | (algorithm* adj2 (pharmacotherap* or chemotherap* or chemotreatment* or therap* or treatment* or intervention*)).ti,ab,kf,kw. | 9314 |  |
| 70 | 46 or 47 or 48 or 49 or 50 or 51 or 52 or 53 or 54 or 55 or 56 or 57 or 58 or 59 or 60 or 61 or 62 or 63 or 64 or 65 or 66 or 67 or 68 or 69 | 1422311 |  |
| 71 | (systematic adj3 review$).ti,ab,kw. | 164344 |  |
| 72 | 24 or 45 or 70 or 71 | 5190597 |  |
| 73 | 10 and 19 and 72 | 3984 |  |

**Appendix B – Detailed Eligibility Criteria**

- *Population (diagnosis)*: Clinicians should discuss their perspectives of services for people with a diagnosis of ‘personality disorder’ or related symptoms/diagnoses. Related symptoms include repeated self-harm or suicide attempts, complex trauma or complex PTSD, and emotional dysregulation or instability (We acknowledge this list is not comprehensive). The primary diagnosis or focus of treatment of the sample should be personality disorder or similar. Treatments that are primarily for other conditions are not included. This excludes qualitative studies of specialist treatment services for other diagnoses including substance misuse conditions.
- *Population (clinicians)*: Relevant stakeholder groups include psychiatrists, psychiatric nurses, psychologists, general practitioners, peer support workers, service managers, commissioners and other professionals who work with people who have received a diagnosis of “personality disorder” or have related symptoms or needs.
- *Settings*: Community services providing mental health support for people who have been diagnosed with a “personality disorder” or experience symptoms or needs which have been associated with “personality disorder”.

Services can be a specialist ‘personality disorder’ service or a generic mental health/primary care/community service, as long as there is some reference to care for people with a diagnosis of personality disorder or related needs. For example, an eligible paper could describe initiatives within generic mental health/primary care service which focus on better meeting the needs of this group of people or describe the experiences of this group of people receiving generic services.

Services for young people will be excluded unless they are transitioning to adult services. For example, qualitative data of people’s experiences of CAMHS treatment per se will be excluded. But data from people’s experiences of moving from CAMHS to adult services will be included.

Hospital services, such as day hospital treatments, are included provided participants still reside in the community through the course of treatment. Participants recruited from inpatient settings may be included provided the treatment being evaluated is a community/outpatient treatment to which they have transitioned. Otherwise, inpatient and crisis services are excluded, as are forensic services and services specifically for offenders.

- *Limits:* Studies published since 2003 will be eligible. 2003 marked a change in UK policy for “personality disorder” services with the publication of implementation guidance for the development of services 'Personality Disorder: No longer a diagnosis of exclusion'. (NIMHE, 2003)
- *Design*: Data should be qualitative and analysed using a recognised qualitative method (such as thematic analysis). Written data from questionnaires can be included if the data are analysed using a recognised qualitative method.

Data from trials and other quantitative studies will be included if the study is naturalistic (i.e. offers a similar delivery/support to a community mental health team or specialist service, e.g. a good quality DBT treatment delivered by mental health practitioners embedded in a mental health service).

- Studies or reports that are available in English or can be translated into English by the research team are eligible – and both published peer-reviewed articles and grey literature are eligible. Grey literature refers to anything which has not been published in a peer reviewed journal.

**References**

National Institute for Mental Health for England. *Personality Disorder: No Longer a Diagnosis of Exclusion. Policy Implementation Guidance for the Development of Services for People with Personality Disorder*. London: NIMH(E); 2003.
